# AI facilitated sperm detection in azoospermic samples for use in ICSI

**DOI:** 10.1101/2023.10.25.23297520

**Authors:** DM. Goss, SA. Vasilescu, PA. Vasilescu, S. Cooke, SHK. Kim, GP. Sacks, DK Gardner, ME. Warkiani

**Affiliations:** School of Biomedical Engineering, University of Technology Sydney, Sydney, 2126, Australia; NeoGenix Biosciences Pty Ltd, Sydney, 2126, Australia; IVFAustralia, Sydney, 2015, Australia; University of New South Wales, Sydney, 2052, Australia; Melbourne IVF, Melbourne, 3002, Australia; Institute for Biomedical Materials & Devices (IBMD), University of Technology Sydney, Sydney, 2126, Australia

**Keywords:** AI, azoospermia, ICSI, mTESE, sperm, SSC

## Abstract

**Research question:** Can artificial intelligence (AI) improve efficiency and efficacy of sperm searches in azoospermic samples?

**Design:** This two-phase proof-of-concept study beginning with a training phase using 8 azoospermic patients (>10000 sperm images) to provide a variety of surgically collected samples for sperm morphology and debris variation to train a convolutional neural network to identify sperm. Secondly, side-by-side testing on 2 cohorts, an embryologist versus the AI identifying all sperm in still images (cohort 1, N=4, 2660 sperm) and then a side-by-side test with deployment of the AI model on an ICSI microscope and the embryologist performing a search with and without the aid of the AI (cohort 2, N=4, >1300 sperm). Time taken, accuracy and precision of sperm identification was measured.

**Results:** In cohort 1, the AI model showed improvement in time-taken to identify all sperm per field of view (0.019±0.30 x 10^-5^s versus 36.10±1.18s, P<0.0001) and improved accuracy (91.95±0.81% vs 86.52±1.34%, P<0.001) compared to an embryologist. From a total of 688 sperm in all samples combined, 560 were found by an embryologist and 611 were found by the AI in <1000^th^ of the time. In cohort 2, the AI-aided embryologist took significantly less time per droplet (98.90±3.19s vs 168.7±7.84s, P<0.0001) and found 1396 sperm, while 1274 were found without AI, although no significant difference was observed.

**Conclusions:** AI-powered image analysis has the potential for seamless integration into laboratory workflows, and to reduce time to identify and isolate sperm from surgical sperm samples from hours to minutes, thus increasing success rates from these treatments.

## Introduction

Male infertility is increasing worldwide at an alarming rate, sperm counts declining by 50% over the past 50 years (Levine et al., 2023). 30% human infertility cases are caused solely by male infertility and 50% of cases being attributed to male infertility as a contributing factor (Agarwal et al., 2015). While assisted reproductive technologies (ART) have proven to be effective in treating infertile couples, some forms of male infertility remain difficult to treat. Azoospermia, defined as the absence of spermatozoa in centrifuged semen on at least two occasions, is the most severe form of male infertility, affecting 10-20% of infertile men and 1% of the general male population (Verheyen et al., 2017, Wosnitzer et al., 2014).

Azoospermia can be classified as either obstructive and/or non-obstructive. Obstructive azoospermia (OA) occurs due to obstruction of the reproductive tract and constitutes 40% of azoospermic cases, while non-obstructive azoospermia (NOA) results from either primary, secondary or incomplete/ambiguous testicular failure which compromises sperm production and constitutes 60% of azoospermic cases (Jarow et al., 1989, Wosnitzer, et al., 2014). Patients with OA can attempt for reconstruction (vasovasostomy, vasoepididymostomy or transurethral resection ejaculatory duct) when possible, or surgical sperm collection can be performed from the testis via testicular sperm aspiration (TESA), testicular sperm extraction (TESE) or microdissection testicular sperm extraction (mTESE) or the epididymis Microsurgical epididymal sperm aspiration (MESA) or percutaneous epididymal sperm aspiration (PESA) (Schrepferman et al., 2001) (Flannigan et al., 2017). NOA patients will require sperm extraction from the testis (TESA, TESE or Micro TESE) and surgically collected sperm is then used for ICSI.

The gold-standard for treating NOA patients, is mTESE, with a high sperm retrieval rate of up to 64% in suitable patients operated on (Deruyver et al., 2014, Ramasamy et al., 2005, Schiff et al., 2005). Although these rates seem promising, the current manual examination process to find sperm within tissue recovered from mTESE surgeries is time-consuming and inefficient, typically taking anywhere between 1-6 h of laboratory time, and in some cases even up to 14 h (Mangum et al., 2020, Ramasamy et al., 2011). This extended time is due to the requirement for manual searching through prepared suspensions of testicular tissue with a microscope, before using isolated sperm for ICSI (Figure 1).

**Figure 1.**
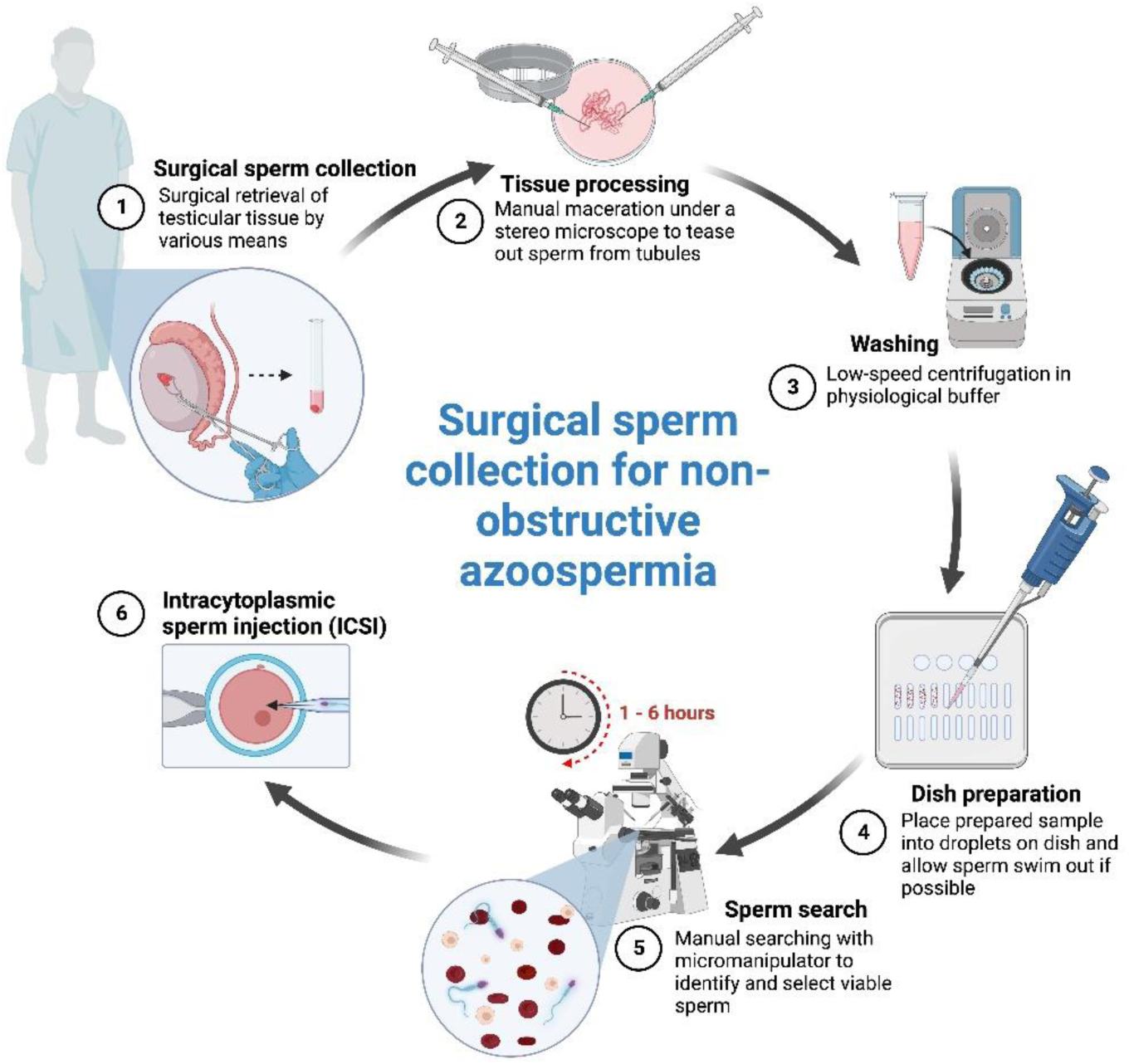
An overview of the non-obstructive azoospermia (NOA) surgical sperm surgery and retrieval process. Tissue from surgery is processed by maceration and washed before being placed in media droplets for manual sperm search which can take up to 6 hours. Viable sperm are then used for intracytoplasmic sperm injection (ICSI).

The outcome of such searching is heavily dependent upon the complexity and contamination of the suspension provided to them by the surgeon. Viable sperm are easily overlooked due to variables such as collateral cell density, resulting in a process that is prone to human error, combined with inexperience and fatigue of lab staff (Ramasamy, et al., 2011). For patients with NOA, if sperm are overlooked due to human error, this could wrongly indicate absolute infertility (Samuel et al., 2016). Similarly, for extended sperm searches in semen as a diagnostic or as a last check in the ejaculate before surgery, failure to identify sperm present could striate patients into surgery unnecessarily. Furthermore, prolonged sample examination procedures can have adverse effects on the viability of sperm, consequently affecting their potential for fertilization (Ouitrakul et al., 2018). For patients with NOA, a more efficient and higher throughput method capable of locating and isolating sperm from the suspension would therefore greatly benefit the clinical workflow of assisting severe forms of male infertility.

Panning through surgically collected sperm samples, under a microscope is a form of manual image analysis in which machine learning (ML) and artificial intelligence (AI) has the potential to automate and improve. Therefore, with preliminary works showing promising results (Goss et al., 2023), this study aims to comprehensively assess the use of an assistive convolutional neural network (CNN) AI to identify sperm in complex tissue suspensions in real time was developed and trained (Figure 2). Using a YOLO V8 model, this software works in tandem with an embryologist to instantly identify and alert embryologists to sperm of interest for their assessment from the camera feed mounted into their microscope. The objective of this study was to compare the AI to embryologists without the AI’s aid in terms of time, accuracy, and number of sperm found first using still images in cohort 1, and then in a simulated sperm search with the AI integrated into an ICSI microscope kit for cohort 2, to demonstrate its potential for clinical implementation.

**Figure 2.**
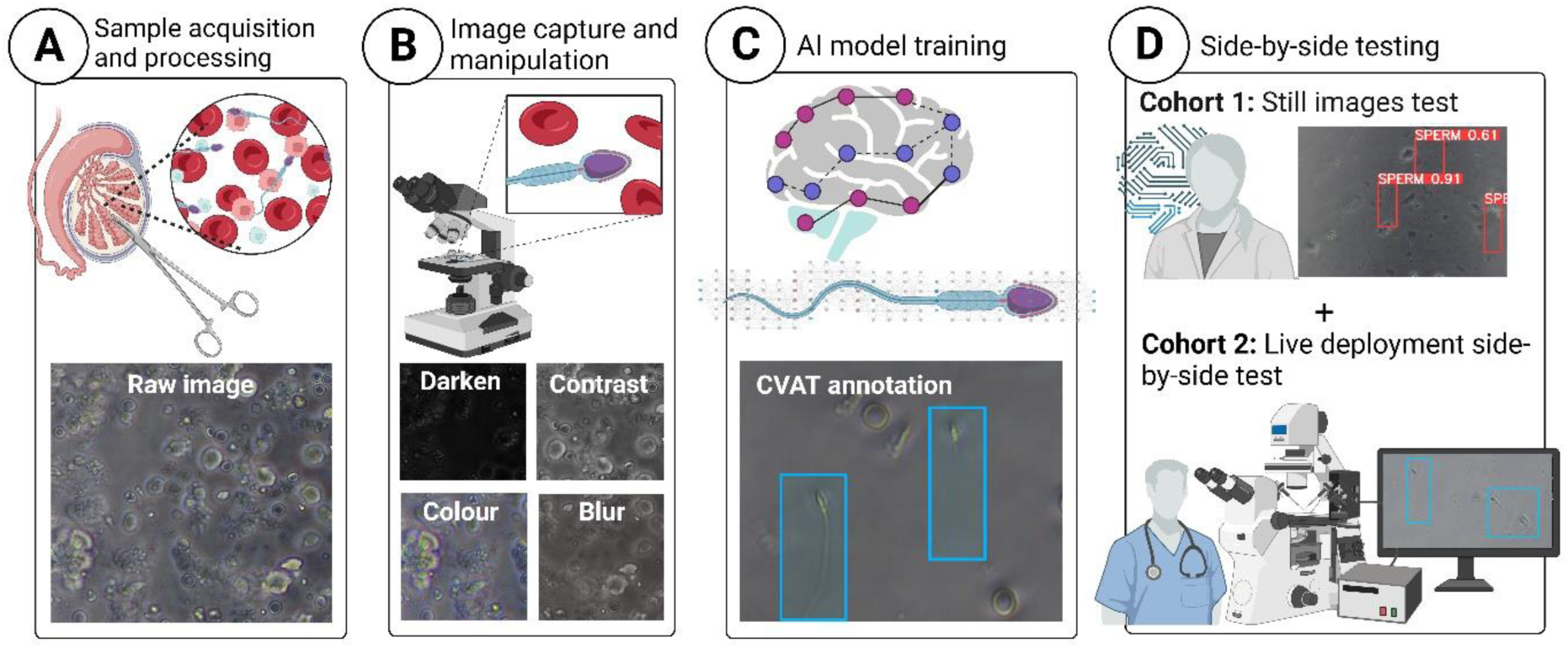
**Overview of the study workflow stages beginning** with (**A**) Sample acquisition and processing from testicular surgery, (**B**) sample image capture and augmentation, (**C**) model training and optimization and (**D**) side-by-side testing comparing AI-based sperm detection and manual assessment by trained embryologists **i**) on still images, as well as **ii**) live deployment comparing searching by an embryologist with and without the aid of the artificial intelligence (AI) model.

## Materials and methods

### Ethical approval

Ethical approval for healthy sperm samples was received from the University of Technology Sydney ethics review board (ETH19-3677), and for the use of discarded testicular tissue samples from the IVFAustralia Human Research Ethics Committee (DG01192) and UTS ethics review board (ETH22-7189).

### Semen collection and processing

Human semen samples were obtained through ejaculation after 2-5 days of sexual abstinence (WHO, 2021). Raw semen samples were left at room temperature for 20 minutes to allow for liquefaction. Samples were centrifuged for 8 minutes at 500 g to separate the sperm pellet from the seminal plasma.

### Cell culture for mock samples for initial training

Red blood cells (RBCs) were obtained from whole blood specimens within three days of collection. Collected blood samples were also resuspended in GMOPS Plus (Vitrolife, Sweden) media. Two sets of mixed cell suspensions were created for microfluidic sperm recovery: a solution of sperm, RBCs, white blood cells (WBCs) and epithelial cells, and a solution of sperm with C2C12 and THP1 cells. All cells were mixed in warmed GMOPS Plus (37°C). Raw semen samples were diluted down to between 1×10^5^ and 1×10^6^ sperm/mL, RBC concentration ranged between 2-15×10^6^ cells/mL (approximated ranges for a mTESE sample), WBCs (purchased from IQ Biosciences, 10×10^6^ cells/mL) were diluted to a concentration between 5×10^5^ and 1×10^6^ cells/mL, and epithelial cells were diluted to a concentration of between 7×10^5^ and 1×10^6^ cells/mL. To add extra complexity, background cells from sperm donors were isolated from 4 donors with high concentrations of background cell populations and cryopreserved until needed. These cells helped simulate the conditions of poor-quality samples with high levels of collateral cell contamination from surgery and for infertile semen samples with high levels of contamination in the ejaculate.

### Testicular biopsy retrieval and processing

Surgical sperm collection was performed in accordance with the routine workflow for each method of sperm collection (mTESE and TESA). Azoospermic patients scheduled for surgical sperm collection for both OA and NOA. Surgical sperm collections were performed under general anesthesia, and the samples were immediately placed in a sterile conical tube containing 1 mL of G-MOPS Plus (37°C) and transported to the IVF laboratory. During mTESE, embryologists search through seminiferous tubules handed to them by the surgeon, with simultaneous further searching by the surgeon for dilated seminiferous tubules. Further samples are then sent to the IVF laboratory for further search before being placed in 1-2 mL of G-MOPS Plus in a sterile petri dish under a stereo microscope, to wash off excess blood from the tissue then moved to a new petri dish with 300 µL of G-MOPS Plus. Tissue was gently teased apart using sterile syringes to release potential sperm from tubules into the surrounding G-MOPS Plus media. The macerated tissue and large pieces were then removed and placed into a separate tube, and the remaining suspension used for the sperm search and treatment. In cases whereby imaging and/or testing was not possible on the nsame day or following day, samples were fixed with 4% formalin to preserve its morphological integrity and prevent any microbial growth until use in the study.

To prepare samples for comparison between AI-enabled sperm search and sperm search by an embryologist in cohort 1, samples that were recorded having no sperm found from clinical searches were spiked with low concentrations of sperm from semen donors. To help create a master count of total sperm in plated samples, spiked sperm were stained with propidium iodide (PI) and washed to remove excess stain before spiking. This was done to help identify the total number of sperm to be found in each sample for comparison with the AI and embryologist performance groups. Samples that had sperm present in the clinics were not spiked with donor semen and preserved in their clinical state for processing.

### Machine Learning Model Development and Training

The training dataset comprises of 540 images, containing 5624 unique sperm instances, duplicated, and augmented generated at least one augmented copy per image, generating over 10 000 sperm to train the identification function. Any image set used to train the model was not used for comparison with the embryologist, although they were prepared similarly. The annotations were made using Computer Vision Annotation Tool (CVAT; Intel, USA). The model used was YOLO v8 with the ‘small’ size architecture configuration with 225 layers and 1,1166,560 parameters to prioritize minimal inference time over potentially greater accuracy from more parameters (Jocher et al., 2023).

Training images were 2456×1842px, JPG with 95% compression. These were resized to 1664×1664px with black fill. 85% of the images were used for training and 15% reserved for validation of the model’s performance after training. Augmentations were applied to duplicates of the images to inflate the dataset and make the trained model more robust to variations in microscope camera images, such as compression artifacts, changing focal length, or lighting and colour variations. A vertical flip was applied to each duplicate image, ensuring it was unique from its source, then with various probability applied 2×2px blur, jpeg compression (60-80%) and multiplicative noise using the Python Albumentations library (Buslaev et al., 2020). Further augmentations are applied by YOLO v8 during the training process, including horizontal flipping, scaling, translation and augments to hue, saturation, and value.

The training setup was restricted to <8GB VRAM. Thus, to maintain a high image resolution we used a small batch size of 4. We trained the model for 300 epochs with a learning rate of 0.01. The stochastic gradient descent optimizer was used with 0.937 momentum and 0.005 weight decay. The trained model is then used to make inferences on unseen, unlabeled images. This results in predictions of where a sperm might be in the image, as well as how confident the model is of each detection as a percentage. We also recorded the time it would take the AI to make predictions on a set of images. This time is dependent on the power of the computer, we opted for a desktop computer with an Intel Core i5-10600K CPU @ 4.10 GHz (6 cores) and an RTX 3070 graphics card.

To evaluate the performance of the model and compared to that of the clinician, we used the model to annotate the same images as are assessed by an embryologist. These annotations are then compared to a ground truth of verified labels to attain a metric of performance including precision and accuracy. Precision is a measure of how many of the model’s detections are correct and accuracy a measure of how many of the sperm in an image the model finds. Detections with significant overlap (>40% Intersection of Union) with confirmed sperm were counted as positive detections and those without as negatives. Detections bordering the edge of an image are often cutoff and lack enough information to distinguish them as either positive or negative, thus any detections within 2px of the edge of the image were omitted.

Proof-of-principle testing was first performed on mock samples containing mixtures of sperm, RBCs, WBCs and epithelial cells from cell culture media. Once the model’s ability to identify sperm was confirmed, clinically obtained testicular tissue samples were used for comparative performance evaluation. Samples were plated in a similar manner to a clinical sperm search, 10 long drops 2-3 mm in length under OVOIL (Vitrolife, Sweden) in an ICSI dish (Vitrolife, Sweden) and imaged.

Side-by-side testing was split into two cohorts both using immotile sperm for the ability to standardize sperm special detection. The first cohort was performed on fixed samples at UTS research laboratories where the total number of sperm in each plated dish was pre-counted as described above, to assess the accuracy, time per field of view (FOV), precision, and number of sperm found. Still images of the droplets (200X) magnification were fed through the AI while embryologists assessed the same images manually and recorded their assessment time per FOV. Embryologists used CVAT to log the location of sperm they identified and the annotated images from the AI were also uploaded into CVAT for comparison. Each group was kept blind to the total count and location of sperm.

The second cohort, to better simulate real-time clinical deployment, a side-by-side test of the AI comparing the performance of an embryologist with and without the AI was performed. After plating a surgical sperm sample into an ICSI dish (Vitrolife, Sweden), the embryologists recorded the number of sperm they found per droplet for each tissue sample they processed with (see supplementary video 1) and without AI, as well as the time taken to complete their assessment. Dishes were blinded to the embryologist and re-ordered to prevent any memory of sperm location by the embryologist when performing each search.

### Statistical Analysis

All statistical analyses were performed using GraphPad Prism 9.0 (GraphPad Software). Normal distribution was assessed using the Shapiro-Wilk Test. The statistical significance of the differences between groups were tested using the two-tailed unpaired Student’s t-test or Mann– Whitney U test if the data were not normally distributed. Two-way analysis of variance to assess the effects of the counting method and sample were performed. P < 0.05 was considered statistically significant and means are expressed with Standard Error of the Mean (SEM) as a measure of sample mean estimates.

## Results

In the first cohort of this study (N=4 patients, 512 still images and 2660 sperm to be found), the AI model showed dramatic improvement in time taken to identify sperm per FOV, improved accuracy in identifying sperm as well as a high level of precision (Figure 3A-C). The AI was able to identify all sperm within each field of view (FOV) in significantly less time compared to the trained embryologist, with a duration of 0.019 ± 0.3 x 10^-5^s versus 36.10 ± 1.18s, respectively (P < 0.0001; Figure 3A). This represents an approximate 99.95% reduction in time per FOV. The AI model demonstrated a significant difference in accuracy compared to the trained embryologist (91.95 ± 0.81% vs 86.52 ± 1.34%, P < 0.001; Figure 3B). The model exhibited a precision of 89.58 ± 0.87%, considering the correct identification of sperm and false positives relative to the control count (Figure 3C). In contrast, the embryologist had a precision of 98.18 ± 0.38%. Out of a total of 2660 sperm, the embryologist identified 1937, while the AI model detected 1997 (Table 1).

**Figure 3.**
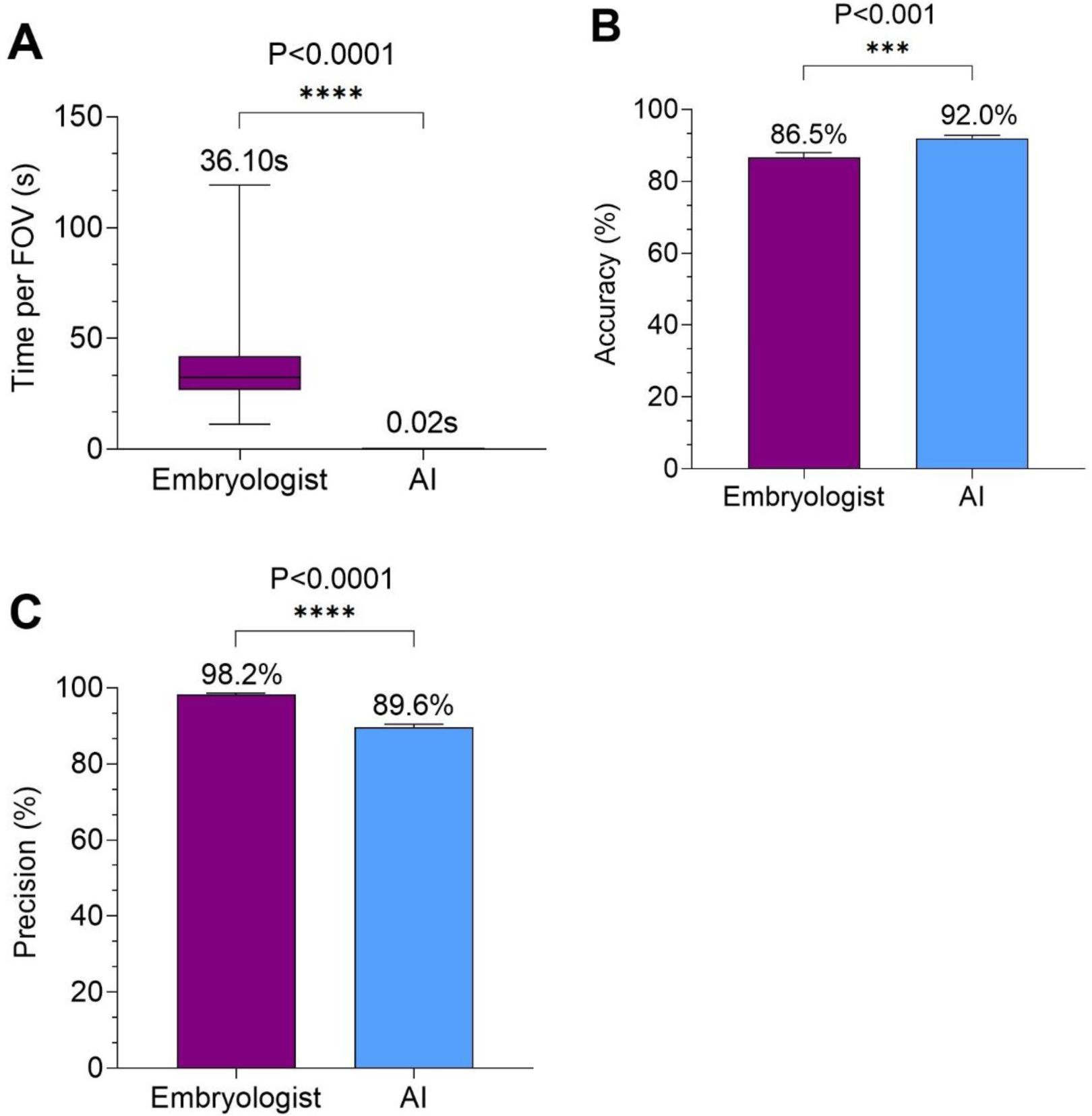
Quantified comparison on still images between the AI and trained embryologists benchmarked against samples with a known number of sperm (cohort 1). A) Time per FOV, B) Accuracy and C) Precision. AI, artificial intelligence; FOV, field of view.

In the second cohort of this study (N=4 patients, 40 media droplets with sample and > 1400 sperm to be found), a simulated deployment of the AI was performed in a research laboratory whereby the AI was used as an assistive tool to guide embryologists to identify sperm on a ICSI microscope kit (see supplementary video 1). Like cohort 1, the AI-assisted embryologist outperformed the individual assessment of an embryologist across all 4 samples. The embryologist using the AI took significantly less time to find all sperm per droplet (98.9 ± 3.19 vs 168.7 ± 7.84, P < 0.0001) and found a total of 1396 sperm while they found 1274 without the use of the AI (Figure 4A; Table 1). There was no significant difference in the number of sperm found per droplet for the embryologist using AI versus without the use of AI although a slight trend of more sperm found, consistently, was observed (34.9 ± 3.23 vs 31.85 ± 3.09 sperm respectively; Figure 4B).

**Figure 4.**
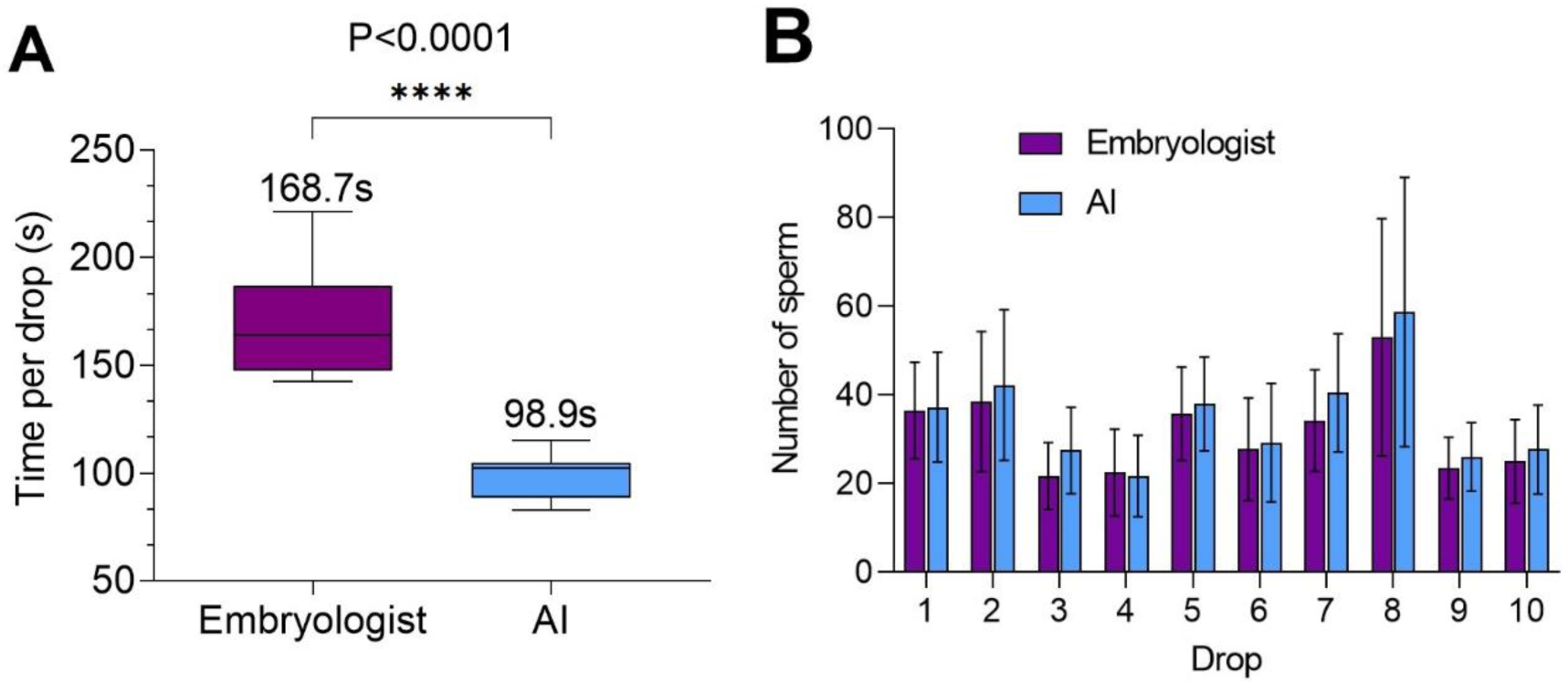
Side-by-side comparison between trained embryologists with and without the use of AI (cohort 2). (**A**) Time taken per droplet and (**B**) number of sperm identified per droplet for each group. AI, artificial intelligence; s, seconds

## Discussion

AI image analysis can identify sperm faster and more accurately than an embryologist in still images and significantly faster in a simulated sperm search scenario when integrated into an ICSI microscope. This is the first known application of ML AI for surgical sperm searches for the clinical treatment of azoospermia and results in a streamlining of a historical laborious process. ML is an algorithmic method of data analysis whereby a predictive model is trained to recognize patterns and associations from input data (Bannach-Brown et al., 2019). Supervised ML models can be trained on labelled images and/or video to understand how to predict the labels of unseen data. CNN algorithms are a type of deep-learning model that attempts to simply replicate the human visual cortex with a simulated network of connected neuron layers (neural network) that, through iterative training, transforms input data into the desired output labels. There have been considerable studies on the utility of machine learning and AI-based image analysis on the selection of embryos for prediction of euploidy status, implantation potential and incidence of miscarriage (Barnes et al., 2023, Diakiw et al., 2022, Duval et al., 2023, Hariharan et al., 2019, Tran et al., 2018, VerMilyea et al., 2020). Studies have also proven the application of ML in the selection and assessment of sperm for use in ICSI by tacking sperm correlated with better quality blastocysts (Joshi et al., 2023, Mendizabal-Ruiz et al., 2022). Furthermore, studies using images of sperm having been labelled as normal or abnormally shaped by a professional or stained for DNA integrity; given a sufficient volume and variety of these labelled images, ML models have been trained to label the morphology of predict DNA fragmentation of new, unseen, images of sperm (McCallum et al., 2019, Wang et al., 2019). Where the CNNs, commonly referred to as AI have largely looked at sperm in a clear environment, we have applied a CNN on complex, processed tissues from testicular sperm retrieval procedures and implemented it in a live video feed to real time identification of sperm for ICSI.

The application of a computer vision-based ML model to identify sperm in real-time during sperm searches outperforms embryologists’ manual searching significantly using still images in time taken, accuracy and sperm count. The biggest noticeable difference is in the time reduction, where image analysis is almost instant (0.02 s per field of view) but does not consider clinical tasks such as dish setup, panning and magnification change, and collection of identified sperm. Significantly lower time taken to identify sperm per FOV (Figure 3A), higher accuracy (Figure 3B), and an increase in the total number of sperm found show clear superiority of AI image analysis compared to the eyes and focus of trained embryologists (Table 1). Although the AI had a lower precision value than embryologists in the first cohort (Figure 3C), it is worth noting that this is a result of the annotation approach taken when training the AI. In the second cohort, testicular tissue samples with supplemented sperm (for better quantification of efficacy) were searched by an embryologist in mock ICSI dishes to better simulate a clinical sperm search on an ICSI kit with and without the aid of the AI (see supplementary video 1), it was determined that the AI reduced the time taken to identify all sperm in the droplet by around 50% (Figure 4A), with no drop in number of sperm identified per drop (Figure 4B) and a higher total number of sperm identified (Table 1).

The role of the AI is not to replace an embryologist, but to be a guide towards sperm of interest, leaving the embryologist to make the final determination on the suitability of a sperm for ICSI. AI can negate the biological limits of human error and observation as well as the effects of fatigue which have long been a limiting factor to extended sperm searches of heterogenous samples obtained via surgical sperm collection. It is important to remember however, the AI is limited to detection within the manually directed field of view, thus if the embryologist overlooked an area in the sample, the AI will not be able to detect sperm without having it within view.

Using an exhaustively trained image analysis model to identify sperm based on tens of thousands of sperm images has clinical utility in directing an embryologist’s attention to what the AI deems may be of interest and can thus drastically reduce the time taken or manual extended sperm searches when integrated to a micromanipulator microscope. The CNN trained in this study is designed to cater for multiple clinics which may have different microscopes, light environments, filters, and cameras. These environmental and equipment factors may affect the performance of the AI and have thus been catered for. The image augmentations used to train the AI on, provide as much variety as possible and with carefully considered image augmentations such blur, colour variations, focus changes, image saturation, and colour balance changes make the model resilient and consistent (Figure 2). The model was also trained using both epididymal and testicular sperm to broaden the sample dataset empowering the AI to broaden target sperm prompting. The model can also identify sperm with the entire range of motility, from immotile to hyperactivated, and adjusts and adapts to magnification change and panning in real-time (see supplementary video 2).

This study was performed solely on immotile sperm for the most accurate quantification for spatially identifying and locating sperm, although the AI identifies motile sperm very well, a clinical deployment will better prove the clinical utility model. This proof-of-concept study demonstrates the potential for AI-assisted sperm searches, both in semen for extended sperm searches and testicular tissue but. While the results of this study are promising, continuing to improve the core data set and image variety will make the model more robust and adoptable for clinics with significantly different microscope arrangements as well as achieving a higher level of accuracy. The limitation of a simulated sperm search using an ICSI workstation with and without the use of the AI, using samples with spiked in sperm, do not consider the time spent confirming the locations of sperm in the field of view during panning (as to not recount or miss sperm), this is a drawback of the testing method and can contribute to the lower difference in time taken per method in cohort 2. Therefore, a robust clinical deployment study has been planned for consenting in-treatment patients whereby embryologists can perform sperm searches with the aid of the AI model. Furthermore, there is potential for the expansion of this AI to include motility and morphological assessments of identified sperm to aid in the choice of sperm for insemination when sperm outnumbers the number of oocytes suitable for injection.

In conclusion, azoospermia affects 10% of infertile men, with NOA, the most severe form, constituting 60% of these cases (Verheyen, et al., 2017). The current approaches to recover sperm from men who undergo surgery from this condition are antiquated and potentially detrimental to the quality of the sperm found. In this study, we have successfully demonstrated a proof-of-concept application of an AI image analysis model to drastically reduce sperm search time in testicular tissue samples in simulated clinical sperm searches. When applying the AI to a simulated real-time search workflow, a 50% reduction in time taken to identify sperm has been demonstrated. Applying this approach with further development and ergonomic optimization, we believe it can result in a standardized and more efficient workflow, greatly improving the current processing procedure of all surgically retrieved samples and azoospermic ejaculates by increasing access to treatment for azoospermia and reducing staff time required, as well as increasing sample coverage to ultimately increase chances of finding sperm.

## Data Availability

All data produced in the present study are available upon reasonable request to the authors

## Authors roles

DMG, SAV, PAV, SC, SHK, DKG and MEW designed and conceptualized the study. DMG, SAV and PAV were responsible for all data acquisition. PV and SAV designed and trained the AI model. DMG, SHK and SAV facilitated clinical sample acquisition. DMG, SAV and PAV drafted the manuscript and all authors critically revised, finally approved and all agree to be accountable for academic integrity and accuracy of the research.

## Acknowledgments

We would like to acknowledge the support and facilitation provided by IVFAustralia Eastern Suburbs doctors, providing access to patients undergoing treatment for severe male factor infertility and technical support and input from the embryology team. M.E.W. would like to acknowledge the support of the Cancer Institute New South Wales through the Career Development Fellowship (2021/CDF1148).

## References

Agarwal A, Mulgund A, Hamada A, Chyatte MR, 2015. A unique view on male infertility around the globe. Reproductive biology and endocrinology. 13, 1–9.

Bannach-Brown A, Przybyła P, Thomas J, Rice AS, Ananiadou S, Liao J, Macleod MR, 2019. Machine learning algorithms for systematic review: reducing workload in a preclinical review of animal studies and reducing human screening error. Systematic reviews. 8, 1–12.

Barnes J, Brendel M, Gao VR, Rajendran S, Kim J, Li Q, Malmsten JE, Sierra JT, Zisimopoulos P, Sigaras A, 2023. A non-invasive artificial intelligence approach for the prediction of human blastocyst ploidy: A retrospective model development and validation study. The Lancet Digital Health. 5, e28–e40.

Buslaev A, Iglovikov VI, Khvedchenya E, Parinov A, Druzhinin M, Kalinin AA, 2020. Albumentations: fast and flexible image augmentations. Information. 11, 125.

Deruyver Y, Vanderschueren D, Van der Aa F, 2014. Outcome of microdissection TESE compared with conventional TESE in non-obstructive azoospermia: a systematic review. Andrology. 2, 20–24.

Diakiw S, Hall J, VerMilyea M, Amin J, Aizpurua J, Giardini L, Briones Y, Lim A, Dakka M, Nguyen T, 2022. Development of an artificial intelligence model for predicting the likelihood of human embryo euploidy based on blastocyst images from multiple imaging systems during IVF. Human Reproduction. 37, 1746–1759.

Duval A, Nogueira D, Dissler N, Maskani Filali M, Delestro Matos F, Chansel-Debordeaux L, Ferrer-Buitrago M, Ferrer E, Antequera V, Ruiz-Jorro M, 2023. A hybrid artificial intelligence model leverages multi-centric clinical data to improve fetal heart rate pregnancy prediction across time-lapse systems. Human Reproduction. 38, 596–608.

Flannigan R, Bach PV, Schlegel PN, 2017. Microdissection testicular sperm extraction. Translational Andrology and Urology. 6, 745.

Goss D, Vasilescu S, Vasilescu P, Sacks G, Gardner D, Warkiani M, 2023. O-136 Artificial intelligence to assist in surgical sperm detection and isolation. Human Reproduction. 38, dead093. 163.

Hariharan R, He P, Meseguer M, Toschi M, Rocha JC, Zaninovic N, Malmsten J, Zhan Q, Hickman C, 2019. Artificial intelligence assessment of time-lapse images can predict with 77% accuracy whether a human embryo capable of achieving a pregnancy will miscarry. Fertility and Sterility. 112, e38–e39.

Jarow JP, Espeland MA, Lipshultz LI, 1989. Evaluation of the azoospermic patient. The Journal of urology. 142, 62–65.

Jocher G, Chaurasia A, Qiu J. YOLO by Ultralytics. 2023. Ultralytics, GitHub.

Joshi K, Simbulan RK, Rajah AM, Burd G, Gupta S, Behr B, Guarnaccia M, Singh G, 2023. A proof-of-concept prospective study of applying artificial intelligence for sperm selection in the IVF laboratory. Reproductive BioMedicine Online. 103329.

Levine H, Jørgensen N, Martino-Andrade A, Mendiola J, Weksler-Derri D, Jolles M, Pinotti R, Swan SH, 2023. Temporal trends in sperm count: a systematic review and meta-regression analysis of samples collected globally in the 20th and 21st centuries. Human reproduction update. 29, 157–176.

Mangum CL, Patel DP, Jafek AR, Samuel R, Jenkins TG, Aston KI, Gale BK, Hotaling JM, 2020. Towards a better testicular sperm extraction: novel sperm sorting technologies for non-motile sperm extracted by microdissection TESE. Translational Andrology and Urology. 9, S206.

McCallum C, Riordon J, Wang Y, Kong T, You JB, Sanner S, Lagunov A, Hannam TG, Jarvi K, Sinton D, 2019. Deep learning-based selection of human sperm with high DNA integrity. Communications biology. 2, 250.

Mendizabal-Ruiz G, Chavez-Badiola A, Figueroa IA, Nuño VM, Farias AF-S, Valencia-Murilloa R, Drakeley A, Garcia-Sandoval JP, Cohen J, 2022. Computer software (SiD) assisted real-time single sperm selection associated with fertilization and blastocyst formation. Reproductive BioMedicine Online. 45, 703–711.

Ouitrakul S, Sukprasert M, Treetampinich C, Choktanasiri W, Vallibhakara SA-O, Satirapod C, 2018. The Effect of Different Timing after Ejaculation on Sperm Motility and Viability in Semen Analysis at Room Temperature. Journal of the Medical Association of Thailand. 101.

Ramasamy R, Reifsnyder JE, Bryson C, Zaninovic N, Liotta D, Cook C-A, Hariprashad J, Weiss D, Neri Q, Palermo GD, 2011. Role of tissue digestion and extensive sperm search after microdissection testicular sperm extraction. Fertility and sterility. 96, 299–302.

Ramasamy R, Yagan N, Schlegel PN, 2005. Structural and functional changes to the testis after conventional versus microdissection testicular sperm extraction. Urology. 65, 1190–1194.

Samuel R, Badamjav O, Murphy KE, Patel DP, Son J, Gale BK, Carrell DT, Hotaling JM, 2016. Microfluidics: The future of microdissection TESE? Systems Biology in Reproductive Medicine. 62, 161–170.

Schiff JD, Palermo GD, Veeck LL, Goldstein M, Rosenwaks Z, Schlegel PN, 2005. Success of testicular sperm injection and intracytoplasmic sperm injection in men with Klinefelter syndrome. The Journal of Clinical Endocrinology & Metabolism. 90, 6263–6267.

Schrepferman CG, Carson MR, Sparks AE, Sandlow JI, 2001. Need for sperm retrieval and cryopreservation at vasectomy reversal. The Journal of urology. 166, 1787–1789.

Tran A, Cooke S, Illingworth P, Gardner D, 2018. Artificial intelligence as a novel approach for embryo selection. Fertility and Sterility. 110, e430.

Verheyen G, Popovic-Todorovic B, Tournaye H, 2017. Processing and selection of surgically-retrieved sperm for ICSI: a review. Basic and Clinical Andrology. 27, 1–10.

VerMilyea M, Hall J, Diakiw S, Johnston A, Nguyen T, Perugini D, Miller A, Picou A, Murphy A, Perugini M, 2020. Development of an artificial intelligence-based assessment model for prediction of embryo viability using static images captured by optical light microscopy during IVF. Human Reproduction. 35, 770–784.

Wang Y, Riordon J, Kong T, Xu Y, Nguyen B, Zhong J, You JB, Lagunov A, Hannam TG, Jarvi K, 2019. Prediction of DNA integrity from morphological parameters using a single-sperm DNA fragmentation index assay. Advanced Science. 6, 1900712.

WHO. Laboratory manual for the examination and processing of human semen. Sixth edn, 2021. World Health Organisation.

Wosnitzer M, Goldstein M, Hardy MP, 2014. Review of azoospermia. Spermatogenesis. 4, e28218.

